# SARS-CoV-2 PCR and antibody testing for an entire rural community: methods and feasibility of high-throughput testing procedures

**DOI:** 10.1101/2020.05.29.20116426

**Authors:** Ayesha Appa, Gabriel Chamie, Aenor Sawyer, Kimberly Baltzell, Kathryn Dippell, Salu Ribeiro, Elias Duarte, Joanna Vinden, Cliahub Consortium, Jonathan Kramer-Feldman, Shahryar Rahdar, Doug MacIntosh, Katherine Nicholson, Jonathan Im, Diane Havlir, Bryan Greenhouse

## Abstract

High-volume, community-wide ascertainment of SARS-CoV-2 prevalence by PCR and antibody testing was successfully performed using a community-led, drive-through model with strong operational support, well-trained testing units, and an effective technical platform

## Introduction

Since the beginning of the COVID-19 pandemic, impaired access to testing across the United States has limited our understanding of epidemiology and thus limited disease control. With clear evidence of asymptomatic infection^1^ but minimal systematic active surveillance across larger communities, additional efforts to conduct large-scale testing were needed to understand the breadth of COVID-19 disease.

While there have been other efforts to provide drive-through testing (mostly using PCR for symptomatic or exposed individuals)^2–5^, no standard procedures existed to safely and efficiently conduct “pop-up” testing using PCR and antibodies for an entire community. Here, we describe the procedures and methodology associated with safe, high volume comprehensive testing for SARS-CoV-2, the first effort to perform community-wide, universal PCR and antibody testing to our knowledge. By obtaining rapid, comprehensive information about active and past infection, we offer this as one model to augment disease surveillance for rural or suburban populations.

## PRIOR TO TESTING

### Community Mobilization (Patient and Public Involvement)

Support from key stakeholders in the community was crucial to this project’s success; in Bolinas, this project was initiated by and co-led by community members, who served as leaders throughout the planning and operational process. Other key stakeholders included the major community-based health organization, the Department of Public Health, and the Fire Department. Most of these groups, together with study leadership, participated in a virtual Town Hall the week prior to study start to introduce the study to the community and answer questions. Additionally, specific community liaisons engaged people experiencing homelessness, the Latinx community, and home-bound elders to maximize participation. In summary, while each community has distinct needs, we found that an early needs assessment with regard to community mobilization to identify essential community partners was an most important early step.

### Registration & Pre-test Survey

Town residents and local first responders were invited to register online, using a custom interface created on a HIPAA-compliant platform in partnership with study leadership and community liaisons. Residents were directed to begin the process by providing contact information for 2-factor authentication (either phone or email) to ensure security and confirm ability to return results. If they were not able to use the online interface, they could call a local facility, where first-responder volunteers helped people register online.

Participant inclusion in the study was confirmed by providing their zip code or indicating their status as first responders. Participants completed an online consent and survey, which included questions about the household as well as demographics, contact information, travel and movement information, symptoms, and medical history. Each household was scheduled for 15-minute appointments allowing no more than five persons per car, and they received a confirmation of their appointment time by their desired mode of contact (SMS, email). The online and telephonic experience including website, consent, and survey was available in both English and Spanish. On the day of testing, participants were emailed or texted with a brief summary of what to expect during their testing experience.

### Sample Data Management

Robust sample identification was a key aspect to ensuring successful data management, and an important challenge to address in the community-based, “pop-up” context. In accordance with Clinical Laboratory Improvement Amendments (CLIA) regulations specified by the clinical laboratory, our labels contained two identifiers, name and date of birth (see Box 1 for example below). Labels additionally contained a random letter code in human readable and QR code format, to serve as a scannable identifier linking each specimen to a unique participant record in the online database.

Our site was not equipped for on-demand label printing in each lane, so all pre-registered participants had labels pre-printed the morning prior to testing. Each lane contained a packet of alphabetized, pre-printed water-resistant cryo labels. Each participant had 4 identical labels per sheet: two to be used on the two specimen containers, one on a lab requisition sheet, and one spare label. If the participant registered onsite, the administrator either: 1) used an onsite label printer available in some lanes or 2) used a set of labels with a unique barcode but otherwise blank, and handwrote the participant’s name and date of birth on the labels and requisition form.

## Box 1: Sample labels

*A. Preregistered participant*

*B. Empty label for participant registering onsite*

## DURING TEST DAYS

### STAFFING & FLOW

In order to complete testing of more than 1,800 individuals over four days, two tents were set up in a large lot with a lane on either side of each tent, to create four total lanes for testing (Figure 1). Participants’ first interactions were with primarily community volunteers outside the testing area, who then directed participants toward medical staff and volunteers in each lane for testing.

**Figure 1:**
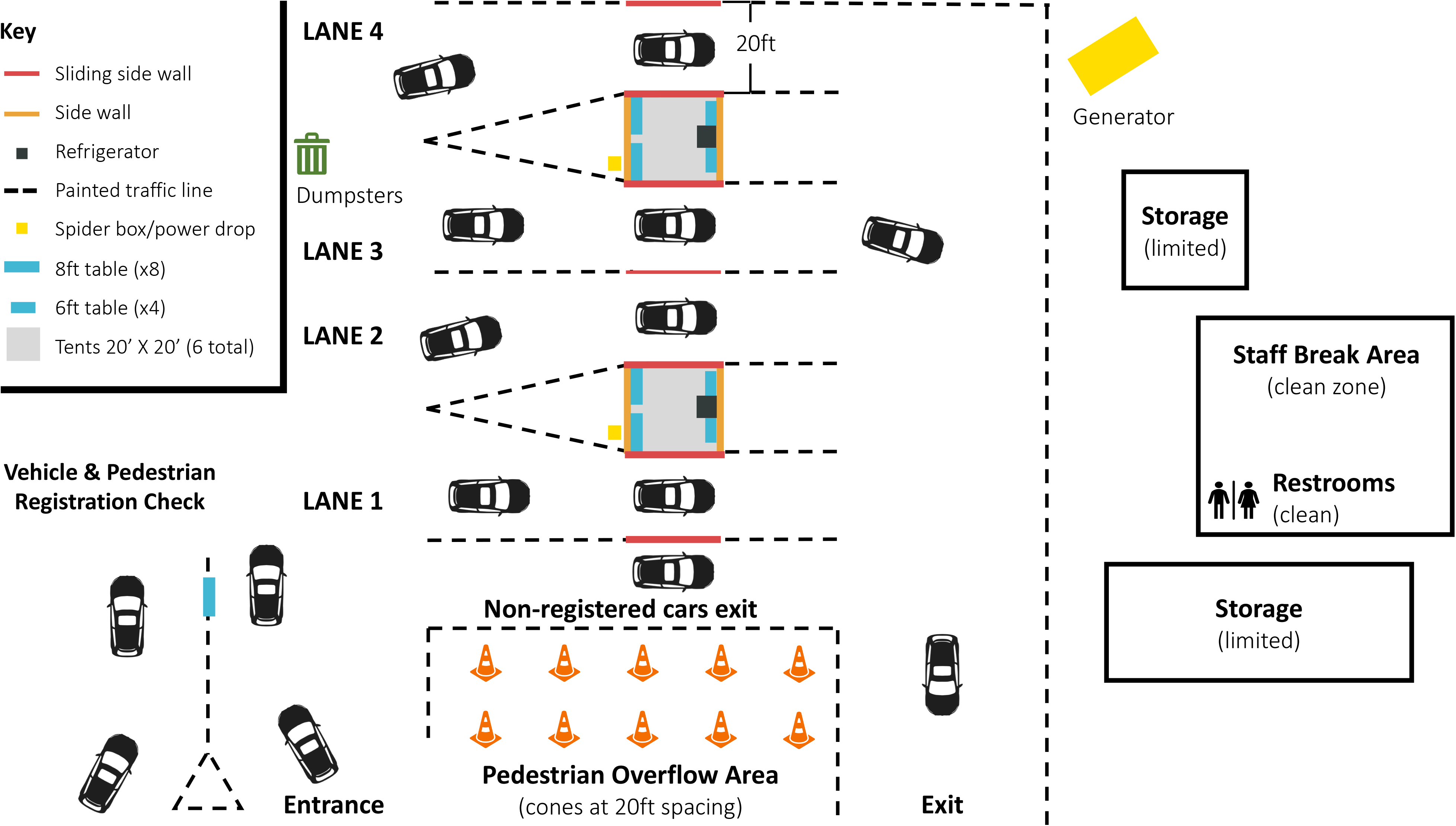

### Site Entry & Lane Triage

To help facilitate entry into the site and prevent interference with regular traffic flow in the area, traffic controllers were stationed at the intersection of the main road. Once cars arrived in the designated area, participants were met by a “greeter” who passed out surgical masks to all participants (Supplemental Table 1). If Spanish-English translation were required, a volunteer translator was engaged at this point (site entry) to help navigate the testing site experience. Local community members volunteered for these three roles, as healthcare experience was not required but local knowledge and a welcoming presence was very helpful.

Next, a “triage greeter” with a tablet confirmed participants’ pre-registration and appointment time and screened for symptoms of COVID-19, including fever, cough, shortness of breath, fatigue, myalgias, anosmia, and dysgeusia. If a participant (or anyone in the vehicle) were symptomatic, they were directed to the specified symptomatic lane. If asymptomatic, the triage greeter directed participants to the shortest lane. Because of the symptom assessment, triage greeters were volunteers in the healthcare field who were comfortable with tablet use. Using the online platform on the tablet, the triage greeter indicated which lane the participant moved into, allowing each lane’s administrator (sitting in the tent) to view the queue of participants in their lane and prepare for their arrival.

## Testing Bay & Tent Staffing/Flow

There was one testing bay per lane, each staffed by 4 people: two dyads of 1) professional phlebotomist who performed testing (“tester”) and 2) a healthcare volunteer (primarily nursing, medical, and pharmacy students) who served as the “test assistant” (Figure 2). Inside the tent, an additional volunteer provided administrative support to each lane. Finally, each tent had two tent supervisors, each of whom was a graduate-level trained nurse, physician, or trained volunteer. As a participant approached the testing bay in any given lane, the test assistant confirmed the participant’s name, date of birth, and whether they had symptoms that day. Without entering the tent themselves, the test assistant in the bay relayed this information back to the administrator in the tent, who logged symptoms and prepared a test kit for each participant.

**Figure 2:**
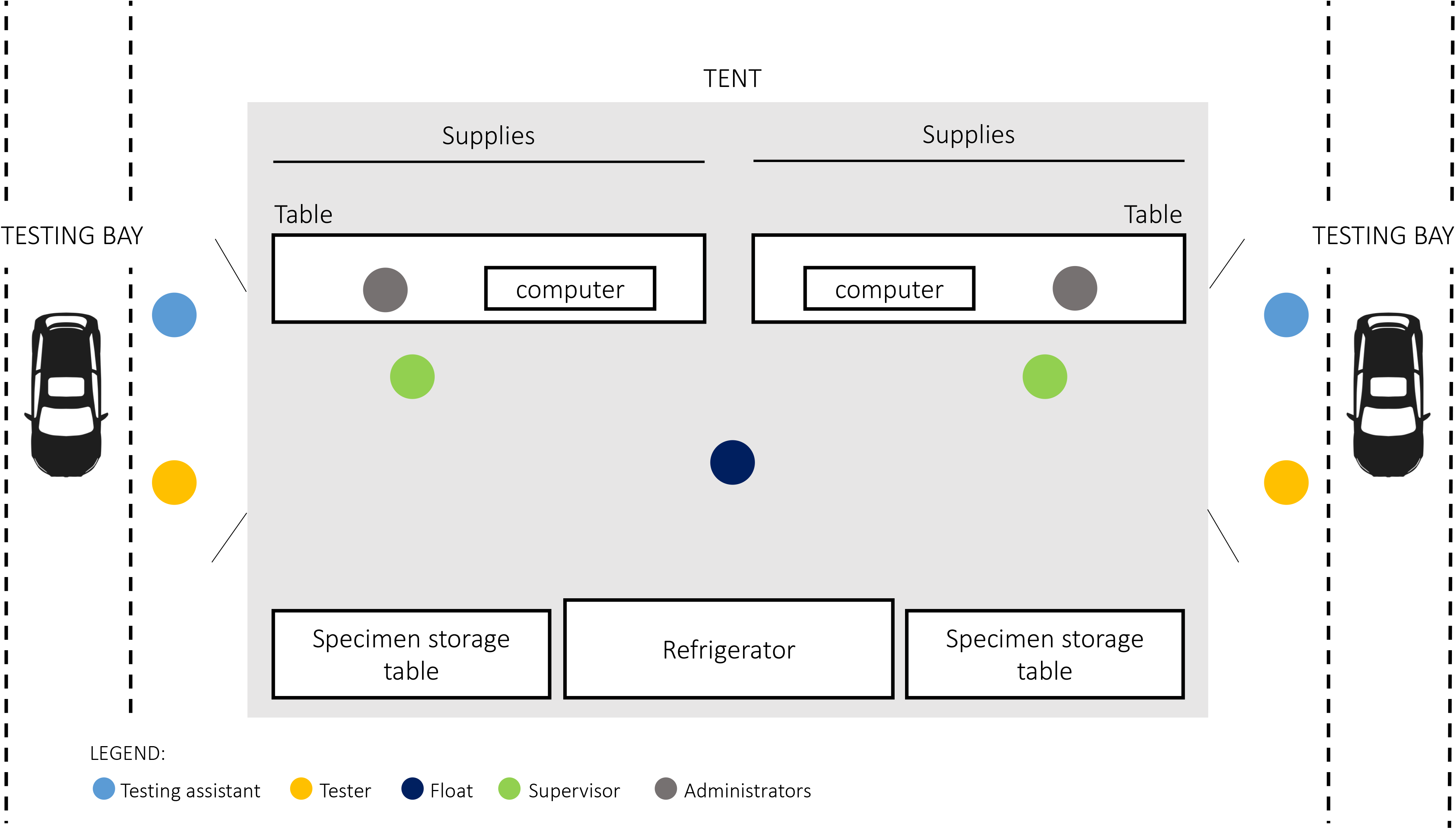

To facilitate throughput, test kits containing all supplies necessary to complete testing in the bay (alcohol wipe, lancet, microtainer, gauze, tongue depressor, swab, viral transport media, biohazard bag) were assembled in advance. The administrator’s role was primarily to locate the appropriate participant labels and afix labels to 1) the microtainer, 2) the viral transport media tube, and 3) requisition sheet. Labeling and test kit preparation was ideally performed in advance of the participant reaching the testing bay, facilitated by the administrator’s ability to view their lane’s queue in the online platform.

In the testing bay, a car pulled into the bay and turned off the engine. If participants arrived on foot or other vehicle, to accommodate those without access to a car, they were seated in a chair in the middle of the lane. The tester explained the procedure, and completed finger stick then oropharyngeal/mid-turbinate swab (see *Test Procedures* for more detail). The test assistant maintained distance from the participant during specimen collection, but was on hand to pass items to the testers. Extra test assistants were trained, with additional test assistants helping as runners/quality control leads when not working actively in the testing bays. Once a participant had completed testing, the test assistant verbally reported completion to the in-tent administrator, and the administrator noted whether tests were successfully administered and that the label barcode matched the database barcode. The participant exited the lane and testing site.

Each tent was also staffed with two tent supervisors, whose role was to trouble-shoot all activities in the testing bay and tent, including responding to participant questions, and ensuring operations ran efficiently. See Supplemental Table 1 for summary of staffing required per day. Finally, on-site staff were screened with an email-based questionnaire before each day to ensure they did not have symptoms associated with COVID-19 (Appendix 1).

## TEST PROCEDURES

Our testing strategy employed both blood collection for antibody testing and upper respiratory tract sampling for PCR testing. With regard to collection of blood, our goal was to maximize community participation by lowering barriers to sampling through use of a finger prick technique (vs. phlebotomy), while collecting enough blood to be sufficient to run quantitative, laboratory-based tests^6^. Please see Appendix 2 for detailed procedures utilized for sample collection.

## Box 2: Test Procedure Pearls

Finger Prick:

- Ensure participants have warm hands prior to finger prick (eg. use of car heaters, hand warmers, hairdryers).
- Use sufficient gauge lancet: we used a 17-gauge push button lancet blade with 2mm depth.
- Prick 3rd or 4th digit of non-dominant hand at lateral aspect of fingertip, then immediately supinate hand well below level of heart to collect specimen.

Respiratory Sampling:

- Instruct masked participants to move their mask down to their chin for oropharyngeal collection, then replace back over mouth (but not nose) for mid-turbinate collection.

## PPE Requirements

We constructed personal protective equipment (PPE) requirements using the following framework adapted from World Health Organization guiding principles: 1) consider the type of contact with participants, 2) incorporate transmission dynamics and environmental factors pertinent to the testing site, and 3) utilize stewardship and appropriate PPE re-use when possible^7^.

For each role, we defined the degree of contact with participants and whether possible to adhere to physical distancing while performing the role. For example, the tester role involved physical contact with participants’ hands and proximity to unmasked participants during oropharyngeal and midturbinate specimen collection that may lead to sneeze or cough. As follows, PPE requirements for this role were the most stringent, including coveralls (or gown), gloves, respirator, and face shield. Conversely, the test assistant did not have physical contact with participants nor were they in close proximity during specimen collection but did talk with participants prior to testing and were also handling specimens after collection. As such, the recommended PPE for this role was a surgical mask, face shield, and gloves. Please see Supplemental Table 1 for detailed PPE recommendations for all roles. When considering generalizability to other testing approaches, the most important consideration should be given to the movements and participant interactions involved in each role with PPE recommendations based on associated exposure risk.

Finally, with regard to PPE reuse, we modeled our guidelines after our medical center and CDC guidelines to minimize waste of materials (Supplemental Table 2)^8 9^. In brief, gowns and gloves were never reused, but face shields and masks (either surgical or respirators) were safely removed, cleaned and stored for reuse throughout the day.

## Post-Testing

At end of each testing day, blood samples in microtainers were stored upright in small cardboard specimen boxes, and viral transport media in biohazard bags was ideally stored upright as well. Specimens were transported to the lab each evening for accessioning overnight. Participants were counseled to expect PCR results within 3–7 days and antibody results within 4–6 weeks. Study staff planned to call each participant with a positive PCR result and direct their results to the Department of Public Health. Additionally, both positive and negative results were delivered via the same online platform through which participants registered. Participants received an SMS or email with a code that allowed them to login to view their results. Alternatively, they had the option of calling a hotline for additional support.

## Testing Site Throughput

In total, 1,840 participants were tested over 4 days using this 4-lane drive-through or walk-up model. Seven participants received home-based testing on a supplemental 5th and final day of testing, to total 1,847 participants overall. Fewer participants were scheduled on the first day of testing to allow for study staff and volunteer acclimation to their roles. On Day 1, the fewest number of participants were tested (n = 338 participants), increasing to the highest number tested on Day 4 (n = 571 participants). Notably, performance on Days 2–4 reflects staffing described above, whereas there were fewer personnel available to staff the testing bays on Day 1.

Figure 3 depicts the number of participants tested per hour across all lanes for each of the four days of onsite testing. When including the hours during which time the testing site was fully open for appointments (9am – 5pm), the median number of participants tested per hour onsite was 57 (interquartile range 47–67). The participants included in this analysis were 1,801 participants with time-stamped checkouts (compared to the total of 1,847), slightly underestimating actual throughput.

**Figure 3:**
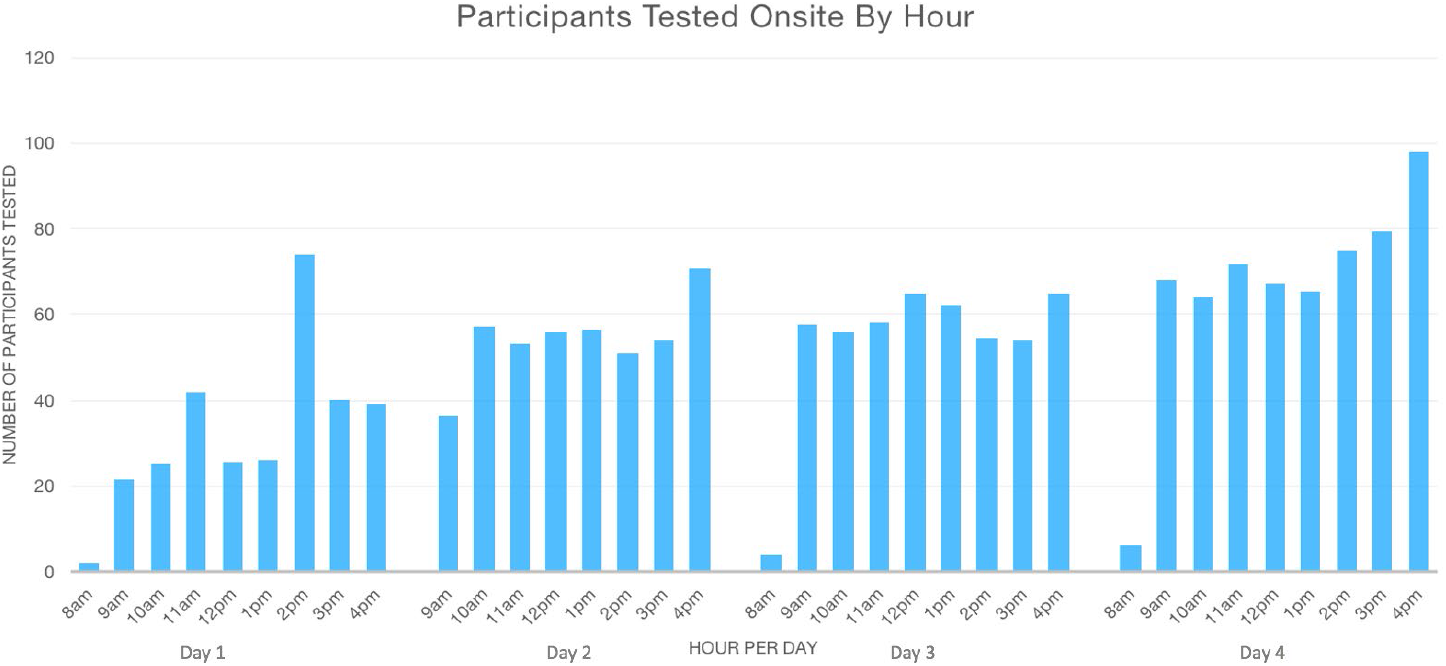

## Identified Areas for Improvement

While this was overall a successful endeavor, we faced a number of challenges that others may improve upon in the future. Our labeling system largely worked well when participants were preregistered, but when participants registered onsite or identified an error in their name or date of birth, handwriting labels was both time-consuming and error prone. Possible improvements include using an onsite label printer along with a barcode scanner in each lane to automate this process as much as possible.

Another challenge was verification of participant identifiers when all parties were wearing masks and maintaining physical distancing. While tent supervisors emphasized ongoing closed-loop communication between team and participants, alternative strategies to verbal communication may offer an improvement. For example, test assistants could use a small white board to write participant identifiers and visually confirm these details with the participant and the administrator.

Finally, despite quality control measures, a small number of viral transport media tubes leaked material upon receipt in the lab. As such, we recommend using tightly sealing vials, inspecting vials prior to testing and upright storage of samples in individual biohazard bags.

An additional suggestion would be to test the entire process in advance of testing roll out, from onsite registration to sample collection to lab reporting. Given the speed with which our efforts were planned, we were limited to testing of individual pieces of the protocol with a final “dress rehearsal” conducted just prior to opening. This issue could also be mitigated by having an experienced team performing the same operations in other locations. That said, our results demonstrate that even in face of rapid planning and new operational system development, we were able to successfully exceed our goals for testing.

## Conclusions

In summary, high-volume, community-wide ascertainment of SARS-CoV-2 prevalence by PCR and antibody testing was feasible and could be performed successfully when conducted in a community-led, drive-through model, with minimal start up time. This operational model may be generalizable to those conducting any sort of high-throughput testing for SARS-CoV-2, regardless of sampling methodology.

## Key Messages

1. High-volume, community-wide ascertainment of SARS-CoV-2 prevalence by PCR and antibody testing could be conducted successfully in a “pop-up” manner, using a multi-lane, drive-through model.
2. Early identification of key community partners for all phases of the project was critical to its success.
3. Robust data management, including contingencies for onsite registration, is a crucial part of early preparation.
4. Proficient testing units, with multiple layers of operational support, can perform increasingly efficient respiratory and fingerstick blood sampling over time.

## Data Availability

This manuscript does not contain patient-level data.

## Contributors, Sources & Acknowledgements

This article was created to share processes, procedures, and lessons learned by our group in creating a novel method of community-based testing that we hope will prove useful to others considering expanding access to SARS-CoV-2 testing. None of the authors have potential conflicts of interest or competing interests to disclose, and author attribution has been included in submission process. We would also like to acknowledge Andrew Kobylinkski for contributing his considerable technical expertise to this project.

## IRB Approval

The aforementioned procedures were submitted to the University of California San Francisco institutional review board for approval, and the study was deemed public health surveillance not requiring IRB oversight [IRB number 20-30636].

